# Automatic identification of risk factors for SARS-CoV-2 positivity and severe clinical outcomes of COVID-19 using Data Mining and Natural Language Processing

**DOI:** 10.1101/2021.03.25.21254314

**Authors:** Verena Schöning, Evangelia Liakoni, Jürgen Drewe, Felix Hammann

## Abstract

**Objectives:** Several risk factors have been identified for severe clinical outcomes of COVID-19 caused by SARS-CoV-2. Some can be found in structured data of patients’ Electronic Health Records. Others are included as unstructured free-text, and thus cannot be easily detected automatically. We propose an automated real-time detection of risk factors using a combination of data mining and Natural Language Processing (NLP).

**Material and methods:** Patients were categorized as negative or positive for SARS-CoV-2, and according to disease severity (severe or non-severe COVID-19). Comorbidities were identified in the unstructured free-text using NLP. Further risk factors were taken from the structured data.

**Results:** 6250 patients were analysed (5664 negative and 586 positive; 461 non-severe and 125 severe). Using NLP, comorbidities, i.e. cardiovascular and pulmonary conditions, diabetes, dementia and cancer, were automatically detected (error rate ≤2%). Old age, male sex, higher BMI, arterial hypertension, chronic heart failure, coronary heart disease, COPD, diabetes, insulin only treatment of diabetic patients, reduced kidney and liver function were risk factors for severe COVID-19. Interestingly, the proportion of diabetic patients using metformin but not insulin was significantly higher in the non-severe COVID-19 cohort (p<0.05).

**Discussion and conclusion:** Our findings were in line with previously reported risk factors for severe COVID-19. NLP in combination with other data mining approaches appears to be a suitable tool for the automated real-time detection of risk factors, which can be a time saving support for risk assessment and triage, especially in patients with long medical histories and multiple comorbidities.

## 1 BACKGROUND

Coronavirus disease 19 (COVID-19) is caused by the severe acute respiratory syndrome coronavirus 2 (SARS-CoV-2). It was first identified in Wuhan, China, in December 2019,(1) and spread globally with rising numbers of cases and deaths.(2)

Several risk factors have been identified for severe COVID-19 disease and need to be taken into consideration when assessing patients in the early stages since they can affect the clinical outcome and are therefore important for patient triage and management decisions. Some of the reported risk factors, such as age,(3, 4) sex,(5, 6) or obesity,(7–10) are included in tabulated form in the electronic health records (EHRs). However, information on other relevant comorbidities such as diabetes,(11–13) cardiac(14–16) and pulmonary diseases,(17, 18) cancer,(19) or dementia(20) is usually included as free-text in the medical history of the EHR and can be time consuming to retrieve. This holds in particular in patients with a long medical history and conditions that might not be included separately in the list of diagnoses (e.g. pre-diabetes). Encoding into ICD (International Classification of Diseases) identifiers – which would be highly suitable for machine processing - is often performed only after discharge or death of the patient and is thus not readily available for the risk evaluation on admission, requires human intervention, and might also only include the main diagnoses relevant to billing purposes.

Here we propose a real-time support for patient triage by automatically identifying relevant risk factors for infection with SARS-CoV-2 and for the development of severe COVID-19 (i.e. need for critical care, mechanical ventilation support, or death of any cause). A combination of data mining and natural language processing (NLP) was used to analyse different parts of the EHRs. Structured (tabulated) administrative, demographic and laboratory data as well as unstructured (free-text) medical history information were processed to identify specific risk factors (age, sex, high BMI, diabetes, cardiovascular disease, pulmonary disorders, dementia, cancer, and decreased kidney and liver function) for SARS-CoV-2 infection or severe courses of COVID-19.(15–17, 21, 22) Furthermore, for diabetic patients, the treatment regimen (untreated, insulin, oral anti-diabetics (OAD) or combination of insulin and OAD) on admission was taken into consideration to classify disease severity.

After development and internal validation, these methods were used in a retrospective analysis of patients seen at the Insel Hospital Group (IHG) in Switzerland, a tertiary hospital network and the biggest health care provider in Switzerland with six locations and about 860’000 patients treated per year. Patients who received testing for SARS-CoV-2 at any point during the ‘first’ and ‘second’ waves of the pandemic in 2020 were included.

## 2 METHODS

### 2.1 Study population

The study was carried out at the IHG. The protocol was approved by the Cantonal Ethics Committee of Bern (Project-ID 2020-00973). We considered all individuals tested for SARS-CoV-2 at the IHG between February 1^st^ through November 16^th^ 2020– covering the ‘first wave’ and part of the ‘second wave’ of COVID-19 in the country, and who did not reject the IHG general research consent. For patients with no registered general research consent status, a waiver of consent was granted by the ethics committee. Patients who objected to the general research consent of the IHG were excluded from the study. Participation in other trials (including COVID-19 related treatment studies) was not an exclusion criterion and was not recorded separately. A reverse-transcriptase polymerase chain reaction (RT-PCR) assay on nasopharyngeal swabs was in use throughout the entire observational period as diagnostic test for SARS-CoV-2 detection. Detailed information on the selection of the study population is provided in **Figure 1**. All patients had been discharged or died by the time of the analysis.

**Figure 1.**
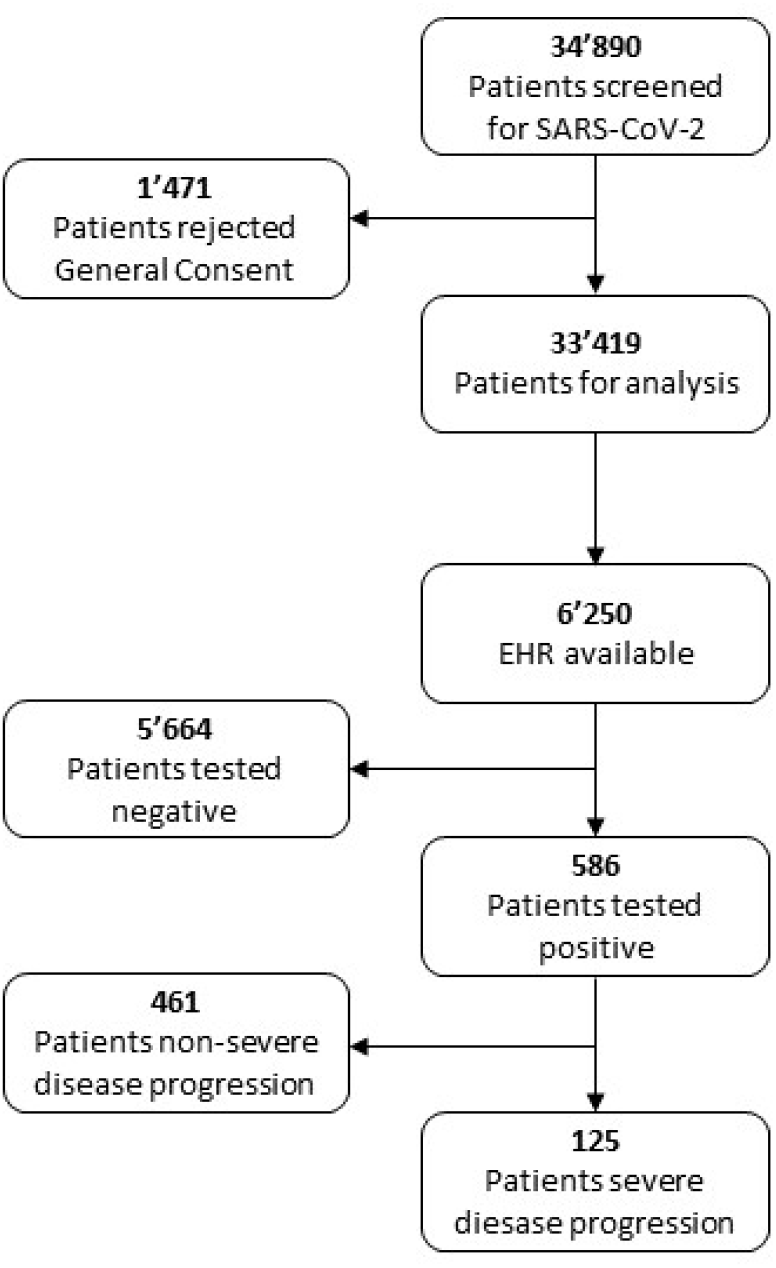
Flowchart of the selection of the patient population included in the study

Patients were classified according to their test result and disease severity, with the worst outcome at any point determining the class:

- *Negative*: Patients who always tested negative for SARS-CoV-2
- *Positive*: Patients who tested positive for SARS-CoV-2 at any point
  - *Non-severe*: Patients who tested positive for SARS-CoV-2, but were neither admitted to the intensive care unit (ICU) nor died of any cause during their hospital stay.
  - *Severe*: Composite outcome for patients who tested positive for SARS-CoV-2 and required ICU admission at any stage during the disease or died of any cause during their hospital stay.

Given that the study was retrospective and observational, sample sizes were dictated by the dynamics of the pandemic in the greater Bern region. Consequently, no formal power calculations were performed *a priori*.

The NLP validation dataset consisted of 138 patients, who were tested positive for SARS-CoV-2 during the first wave in Switzerland (February - August 2020) and whose medical history were available. The EHR was screened manually by two of the authors for the following comorbidities of interest: arterial hypertension, chronic heart failure, atrial fibrillation or flutter, coronary heart disease, asthma, chronic obstructive pulmonary disease (COPD), diabetes (including type I, type II and pre-diabetes), dementia (including mild cognitive impairment), and cancer.(15–17, 21, 22)

#### 2.1.1 Data mining and Natural Language Processing

Age and sex were taken directly from the structured parts of the EHR. Although BMI was also included in the structured parts, some values were not correctly calculated. Therefore, BMI were recalculated using weight and height, which are also documented in the EHR, after removing highly unrealistic values (e.g. weight of 80 kg for a <1 year old child or height of 16.3 cm for an adult), which were probably due to typing errors. Laboratory values indicative for kidney failure (estimated glomerular filtration rate (GFR) according to the Chronic Kidney Disease Epidemiology Collaboration (CKD-EPI) equation) or hepatic dysfunction (alanine aminotransferase (ALAT) and alkaline phosphatase (AP)) were also taken from the structured parts of the EHR. We used the lowest and highest values measured, respectively, in the time frame between three days before and one day after the Sars-CoV-2 test as described in a previous study.(23) Kidney function was categorized into five stages: normal (GFR ≥ 90 mL/min), mild impairment (GFR 60-89 mL/min), moderate impairment (GFR 30-59 mL/min), severe impairment (GFR 15-29 mL/min) and kidney failure (GFR < 15 mL/min).(24) Liver function was considered as impaired if ALAT > 90 U/L (∼3x ULN (upper limit of normal)) and/or AP > 200 U/L (∼2x ULN).

The comorbidities of interest were identified in the medical history, which is the unstructured, free-text part of the EHR. In a first step, we merged the information of different time points into a single text file per patient and removed duplicate sentences. We then read the text files into Python as data corpus, where we performed the actual NLP (**Figure 2**). General data cleaning of these files consisted of converting special characters (e.g. umlauts such as ‘ä’ to ‘ae’), setting the whole text to lower case, and removing dates. Then sentences were tokenized, i.e. the text was separated into single (half-) sentences (tokens). Tokens with less than two characters or information related to family history were removed. As the medical history consisted mainly of catch phrases and half-sentences, we decided against a removal of stop words. For each disease, a specific list of key terms was generated, which consisted of common terms (one or more words) and abbreviations used to describe the diseases in German EHR systems. As there are many different terms for the diverse types of cancer, another approach was chosen for this element. We first scanned all EHRs for words containing cancer-specific terms such as ‘cancer’, ‘carcinoma’, or ‘neoplasia’, and created a key terms list. This list was then appended with other common terms and abbreviations for specific types of cancer, for instance ‘ALL’ for acute lymphoblastic leukaemia. For correction of spelling errors and lemmatization, the key terms list was transformed into a dictionary, where each word only appears once. Words within the medical history were replaced with the corresponding word of the dictionary when the similarity threshold of the Levenshtein distance (minimum number of characters required to transform one word to another) was equal or bigger than 90%. Then, for each token within the medical history, we checked for the presence of the defined key terms. If the key term consisted of more than one word (e.g. ‘arterial hypertension’), the algorithm was set to allow up to three words between the single words of the key term. This further reduced the need for removing stop words.

**Figure 2.**
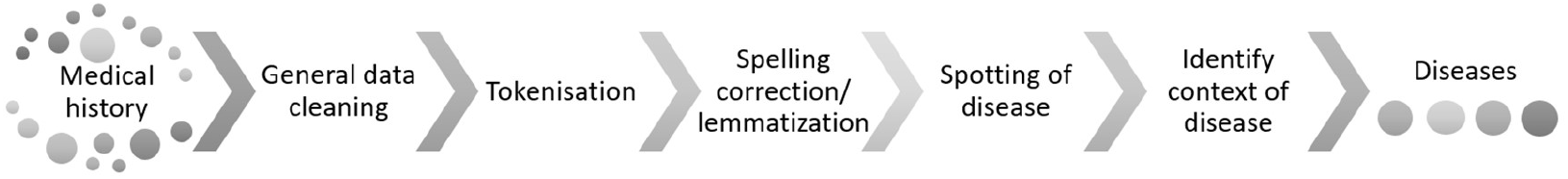
Data preparation steps for the detection of diseases in a free-text medical history

If the key term was present, we determined the context of the key term (affirmation or negation) using a German implementation of *NegEx*.(25) The output of the negation detection was screened, and the negation trigger list appended accordingly where necessary (e.g. ‘non-insulin’). In case of an equal or bigger number of affirmed than negated statements, the patient was tagged as ‘positive’ for the disease. The disease status of each patient as determined by NLP was compared to the manually tagged label. For cases where the automatically detected disease status varied from the manual one, the medical history was screened again, and, if possible, the key term list was corrected accordingly. The NLP detection was considered suitable if the error rate per disease was ≤ 2%.

As the structured medication on admission table was not always filled in, we decided to use a combined approach to identify anti-diabetic treatment: we selected individuals which were identified as diabetes patients by NLP and searched the EHR for key terms of treatment with insulin or OADs. We then combined these NLP results with the medication at admission table. If either the NLP or the medication at admission table showed use of insulin and/or OAD, we tagged the patient as receiving that treatment. If the patient received neither, we categorized the patient as ‘untreated’. For the subgroup analysis of the OAD only cohort, we were only able to use the recorded medication on admission to identify metformin, the only biguanide marketed in Switzerland, as medical histories and diagnosis lists often do not mention the specific OAD used.

Age, sex, BMI, disease status, anti-diabetic treatment and kidney and liver function of all patients with available information were then analysed and stratified according to the cohorts described above.

#### 2.1.2 Software and statistical tests

NLP was performed in Python (version 3.8.5). The text file corpus was generated using the Natural Language Toolkit (*nltk*) package (version 3.5).(26) Steps requiring regular expressions (e.g. text cleaning, sentence tokenization, word replacement) were performed using the *re* package (version 2.2.1). For spelling correction and lemmatization, we used the *FuzzyWuzzy* package (version 0.18.0) and specific, manually generated keyword lists. Detection of negation was performed using *NegEx*(27) in combination with its German implementation.(25)

Data wrangling, analysis and visualization was performed in GNU R (version 4.0.2, R Foundation for Statistical Computing, http://www.R-project.org, Vienna, Austria). Standard statistics, e.g. Shapiro-Wilk test, Chi square, Fisher’s exact test, Wilcoxon rank sum test, were conducted using the *stats* package (version 4.0.2).

Comparisons were performed between a) patients tested negative and positive for SARS-CoV-2 and b) patients with a non-severe and severe clinical manifestation of COVID-19. Statistical significance levels were determined using Wilcoxon rank sum test for non-normally distributed parameters, as confirmed by Shapiro-Wilk test, and the Chi square test or the Fisher’s exact test (sample size ≤ 5) for categorical parameters. A p value of <0.05 was considered statistically significant.

## 3 RESULTS

A total of 6’250 patients tested for SARS-CoV-2 at the IHG during the study period were included and categorized as either SARS-CoV-2 negative (n=5’664) or positive (n=586), and – among the latter-as having a non-severe (n=461) or severe (n=125) clinical outcome. The comorbidities of interest, i.e. arterial hypertension, chronic heart failure, atrial fibrillation or flutter, coronary heart disease, asthma, COPD, diabetes, dementia and cancer, could be automatically detected in the German EHR of the IHG using NLP with an error rate of ≤ 2% compared to the manually tagged EHR. The results of the validation and the key terms used for each disease are shown in the supplementary material (**Table S1** and **Table S2**).

Stratification of patients to the outcome showed that the proportions of patients with old age, male sex, higher BMI, arterial hypertension, or diabetes were significantly higher in the positive SARS-CoV-2 and in the severe COVID-19 cohorts (**Table 1**). Patients with atrial fibrillation or flutter were more likely to be tested negative for SARS-CoV-2, but despite a tendency for more severe clinical manifestations of COVID-19, these results were not significant. The same proportion of patients with coronary heart disease was tested negative and positive for SARS-CoV-2, but with a higher proportion of patients with a severe course.

**Table 1:** Distribution of risk factors as determined by automated analysis of the Electronic Health Records (EHRs)

|  | *Test for SARS-CoV-2* |  |  | *COVID-19 clinical manifestation* |  |  |
| --- | --- | --- | --- | --- | --- | --- |
|  | *negative*<br>*(N = 5'664)* | *positive*<br>*(N = 586)* | *p-value* | *non-severe*<br>*(N = 461)* | *severe*<br>*(N = 125)* | *p-value* |
| **Age (years)** |  |  |  |  |  |  |
| **Median (IQR)** | 63<br>(39, 77) | 65<br>(48, 77) | **0.013<sup>s</sup>** | 62<br>(44, 75) | 69<br>(59, 81) | **<0.002<sup>s</sup>** |
| **Sex** |  |  |  |  |  |  |
| **Female (%)** | 2'650<br>(46.8%) | 246<br>(42.0%) | **0.029\*** | 211<br>(45.8%) | 35<br>(28.0%) | **<0.002\*** |
| **BMI** |  |  |  |  |  |  |
| **Median (IQR)** | 24.6 (21.1,<br>28.7) | 26.5 (23.5,<br>29.7) | **<0.002<sup>s</sup>** | 25.9<br>(23.2,<br>29.4) | 27.9<br>(24.8,<br>30.9) | **0.003<sup>s</sup>** |
| **Diseases** |  |  |  |  |  |  |
| **aHT (%)** | 2,270<br>(40.1%) | 270<br>(46.1%) | **0.006\*** | 192<br>(41.7%) | 78<br>(62.4%) | **<0.002\*** |
| **Chronic heart failure (%)** | 1,591<br>(28.1%) | 140<br>(23.9%) | **0.035\*** | 97<br>(21.0%) | 43<br>(34.4%) | **0.003\*** |
| **Atrial fibrillation or flutter (%)** | 1,025<br>(18.1%) | 84 (14.3%) | **0.027\*** | 59<br>(12.8%) | 25<br>(20.0%) | 0.058* |
| **Coronary heart disease (%)** | 1,052<br>(18.6%) | 92 (15.7%) | 0.098* | 56<br>(12.2%) | 36<br>(28.8%) | **<0.002\*** |
| **Asthma (%)** | 350 (6.2%) | 52 (8.9%) | **0.015\*** | 45 (9.8%) | 7 (5.6%) | 0.203* |
| **COPD (%)** | 612<br>(10.8%) | 45 (7.7%) | **0.023\*** | 28 (6.1%) | 17<br>(13.6%) | **0.009\*** |
| **Diabetes (%)** | 1,129<br>(19.9%) | 155<br>(26.5%) | **<0.002\*** | 111<br>(24.1%) | 44<br>(35.2%) | **0.017\*** |
| **Dementia (%)** | 516 (9.1%) | 55 (9.4%) | 0.885* | 40 (8.7%) | 15<br>(12.0%) | 0.339* |
| **Cancer (%)** | 1,407<br>(24.8%) | 96 (16.4%) | **<0.002\*** | 73<br>(15.8%) | 23<br>(18.4%) | 0.582* |
| **Kidney function** |  |  |  |  |  |  |
| **Normal, GFR >90 mL/min (%)** | 2,131<br>(36.16%) | 196<br>(32.13%) | 0.054 | 163<br>(36.22%) | 33<br>(20.62%) | **< 0.002** |
| **Mild impairment, GFR 60-89 mL/min (%)** | 1,871<br>(31.74%) | 239<br>(39.18%) | **<0.002\*** | 187<br>(41.56%) | 52<br>(32.50%) | 0.055 |
| **Moderate impairment, GFR 30-59 mL/min (%)** | 1,299<br>(22.04%) | 116<br>(19.02%) | 0.095 | 77<br>(17.11%) | 39<br>(24.38%) | 0.058 |

|  | **Test for SARS-CoV-2** |  |  | **COVID-19 clinical manifestation** |  |  |
| --- | --- | --- | --- | --- | --- | --- |
|  | **negative**<br>**(N = 5'664)** | **positive**<br>**(N = 586)** | **p-value** | **non-severe**<br>**(N = 461)** | **severe**<br>**(N = 125)** | **p-value** |
| **Severe impairment, GFR 15-29 mL/min (%)** | 385<br>(6.53%) | 37 (6.07%) | 0.720 | 13<br>(2.89%) | 24<br>(15.00%) | **<0.002\*** |
| **Kidney failure, GFR <15 mL/min (%)** | 208<br>(3.53%) | 22 (3.61%) | 1 | 10<br>(2.22%) | 12<br>(7.50%) | **0.005\*** |
| **Liver function** |  |  |  |  |  |  |
| **Elevated liver enzymes (%)** | 837<br>(16.89%) | 74<br>(13.99%) | 0.101 | 30<br>(9.62%) | 26<br>(26.26%) | **<0.002\*** |
| **Diabetes management** |  |  |  |  |  |  |
| **Untreated (%)** | 336<br>(29.8%) | 38 (24.5%) | 0.210* | 27<br>(24.3%) | 11<br>(25.0%) | 1* |
| **Insulin and OADs (%)** | 258<br>(22.9%) | 43 (27.7%) | 0.213* | 30<br>(27.0%) | 13<br>(29.6%) | 0.907* |
| **Insulin only (%)** | 231<br>(20.5%) | 23 (14.8%) | 0.124* | 10 (9.0%) | 13<br>(29.6%) | **0.003\*** |
| **OAD only (%)** | 304<br>(26.9%) | 51 (32.9%) | 0.143* | 44<br>(39.6%) | 7 (15.9%) | **0.008\*** |
| **Incl. metformin (%)** | - | - | - | 21<br>(18.9%) | 2 (4.6%) | **0.024\*\*** |
| **Excl. metformin (%)** | - | - | - | 23<br>(20.7%) | 5<br>(11.46%) | 0.247** |

Bold numbers indicate significant differences ( $p < 0.05$ ) between compared groups. aHT: arterial hypertension, BMI: body mass index, COPD: chronic obstructive pulmonary disease, GFR: glomerular filtration rate, IQR: interquartile range, OAD: oral anti-diabetics, \$ Wilcoxon rank sum test, \* Chi Square test \*\*Fisher test

Patients suffering from chronic heart failure or COPD were less likely to be tested positive for SARS-CoV-2, but more likely to develop a severe COVID-19 course. In contract, significantly more asthmatic patients were tested positive for SARS-CoV-2 than negative. However, with regards to clinical manifestation, no difference could be observed between the two groups. Cancer patients were less likely to be tested positive for SARS-CoV-2, but no significant difference in severity was observed.

Diabetes treatment was not associated with the risk for tested positive for SARS-CoV-2, but with its clinical consequences. Treatment with OAD only was significantly associated with less cases of a severe outcome, whereas treatment with insulin only showed the opposite effect. A subgroup analysis of the OAD only group showed that this effect might be linked to metformin. No differences were seen for untreated and insulin/OAD co-medicated patients.

Normal kidney function around the time of SARS-CoV-2 testing was associated with benign courses, whereas a severely impaired kidney function of kidney failure was more often seen in severe cases. Additionally, elevated liver enzymes were also seen more often in cases with severe COVID-19.

## 4 DISCUSSION

During the ongoing pandemic, the swift identification of patients at risk for infection and severe courses of COVID-19 is highly important. Manual identification of risk factors is a time-consuming task, especially for patients with co-morbidities, as the respective medical histories might be very long. Using NLP on EHRs poses several challenges, as written clinical text contains abbreviations, acronyms, spelling errors, and nested negations.(28) Furthermore, most available software solutions focus on English language text. We were able to develop a suitable NLP algorithm to automatically detect relevant risk factors for COVID-19 in the medical history of patients at time of admission. The selected key terms allowed for a detection in German EHRs with an error rate of ≤ 2% on the validation dataset and thus can be used for automated real-time detection to support risk assessment and triage at point-of-care.

Old age, male sex, higher BMI, arterial hypertension, and diabetes were identified as risk factors for a positive COVID-19 test result as well as for a more severe clinical outcome. This is in line with observations of other studies and reviews.(29) Old age is generally accepted as a risk factor for severe clinical manifestations, probably due to coexisting age-dependent risk factors (e.g. diabetes, arterial hypertension, coronary heart disease, malnutrition) and remodelling of the immune system (immunosenecence).(3, 4, 30) Sex-specific differences in immune response during COVID-19 might be responsible for the higher vulnerability of males,(5, 31) but also life style (e.g. personal hygiene and smoking)(32) might contribute to this observation. Obesity is not only associated with other comorbidities such as diabetes and arterial hypertension, but also several physiological changes, e.g. chronic inflammation, endocrinal dysfunction, impaired pulmonary perfusion, and immune dysregulation, which may contribute to severe clinical manifestations.(7–10) Furthermore, several comorbidities can often also coexist and a number of two or more comorbidities were a significant factor for a more severe COVID-19 clinical outcome in previous studies.(33)

Diabetes was seen in approximately one third of patients with a severe disease course. This proportion was within the range of other studies.(34, 35) Monotherapy of diabetes before admission in COVID-19 patients with insulin was associated with a higher percentage of severe outcomes (29.6% vs. 9.0%). By contrast, OAD treatment that included metformin was associated with a higher percentage of non-severe disease progression (18.9% vs. 4.6%). This decrease in severity was significant for the metformin group (p < 0.02). OAD treatment that did not include metformin was not significantly associated with disease severity. This is in line with recent clinical reports that metformin treatment of diabetic COVID-19 patients resulted in a reduced mortality rate.(36–38) Metformin acts by activating the AMP-activated protein kinase AMPK,(39) which then phosphorylates angiotensin-converting enzyme 2 (ACE2) causing increase in ACE2 expression.(40) Even though ACE2 serves as molecular target for SARS-CoV-2, it also exhibits protective effects.(41–45) Furthermore, AMPK also inhibits the protein kinase B (AKT)/mammalian target of rapamycin (mTOR) pathway that is essential for viral translation.(46) Finally, metformin, inhibits inflammatory pathways and elevated inflammatory immune responses (cytokine storms).(47)

Possible reasons for the observation that COPD was associated with lower proportion of a positive test for COVID-19, which is also seen in other published data,(48) include a potential effect of smoking, the most predominant risk factor for the development of COPD. Smokers have been reported to be less susceptible to COVID-19.(49, 50) Furthermore, given the common symptoms of COPD such as cough, increased phlegm, shortness of breath, and decreased oxygen levels, it is also possible that due to the similarity in clinical presentation with a SARS-CoV-2 infection might lead to a higher test rate in COPD patients and thus more negative SARS-CoV-2 test results, while respiratory diseases including COPD might be under-diagnosed in some positively tested cases.(33) A possible protective role of regular treatment with inhaled corticosteroids in patients with COPD or asthma was not confirmed by Schultze et al.(51) Despite the often similar symptoms, and the presence of chronic airway inflammation in both COPD and asthma, the generally better outcomes in cases with asthma might be due to the usually younger age of these patients or to the non-reversible structural changes of the airways in case of COPD.

Our study has several limitations. As the IHG is a major hospital centre in the region, patients admitted to the hospital for other reasons were also tested for SARS-CoV-2 if they displayed any symptoms indicative for COVID-19. The patients in the SARS-CoV-2 negative cohort probably have more health-related problems than the general population. This effect is further corroborated as we only analysed patients with available EHRs, and, due to the retrospective nature of the study, had no information on patients tested at the ambulant COVID-19 test centre without being admitted to the IHG as in- or out-patients. This was partially mitigated by including records from the three months preceding diagnosis. Therefore the higher incidence of specific comorbidities in the SARS-CoV-2 negative cohort might represent a selection bias. Additionally, we did not perform a case-controlled study or adjusted for cofounding factors such as smoking or age. Furthermore, with the exception of diabetes and renal function, we did no differentiate between the different stages and severities of the diseases. This issue can be addressed by refining the key terms list in a subsequent study.

The validated error rate for the NLP detection of the different disease is around 2%. Due to this low error rate, the pronounced contrast between significant features, and the large amount of EHRs analysed, it is not expected to affect statistical conclusions. The main focus of the analysis were patients tested positive for SARS-CoV-2, and approximately 25% of these patients were used for the validation of NLP and thus manually screened for the diseases of interest.

## 5 CONCLUSION

In conclusion, we were able to develop a suitable NLP algorithm to automatically detect risk factors for COVID-19 in German EHRs at time of admission with an error rate of ≤ 2%. Our technique should be relative easily transferable to other languages by amending the key terms list and the negation detection accordingly. Use of NLP and data mining can provide a timesaving support for risk assessment and triage at point-of-care, especially in patients with long medical histories and multiple comorbidities. In the future, the same technique could also be used for vaccine prioritization for the automated and time efficient identification of persons at risk.

## Data Availability

The datasets used and/or analyses during the current study are available from the corresponding author on reasonable request.

## 6 DECLARATIONS

### 6.1 Authors’ contributions

FH and VS conceptualized this study. VS and FH performed the data analysis. FH and EL contributed to data extraction. All authors critically revised and approved the final manuscript.

## 6.2 Acknowledgements

We thank Noel Frey, Myoori Wijayasingham, and the Insel Data Science Center for database and infrastructure support.

## 6.3 Competing interest

None.

## 6.4 Funding

None.

## 6.5 Ethics approval and consent to participate

The study was approved by the Cantonal Ethics Committee of Bern (Project-ID 2020-00973). Participants either agreed to a general research consent or, for participants with no registered general research consent status (neither agreement nor rejection), a waiver of consent was granted by the ethics committee.

## 9 SUPPLEMENTARY MATERIAL

**Table S1:**
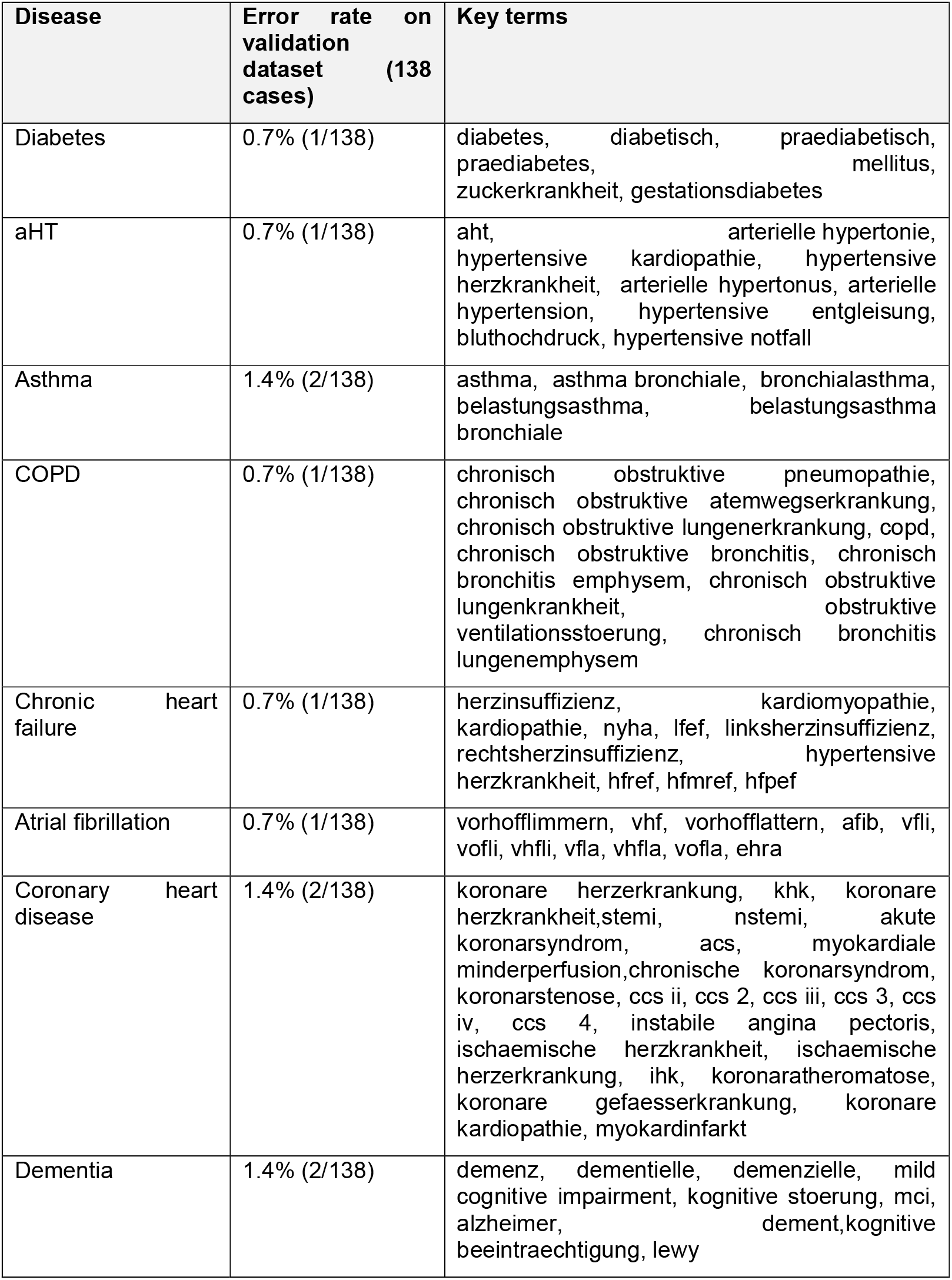
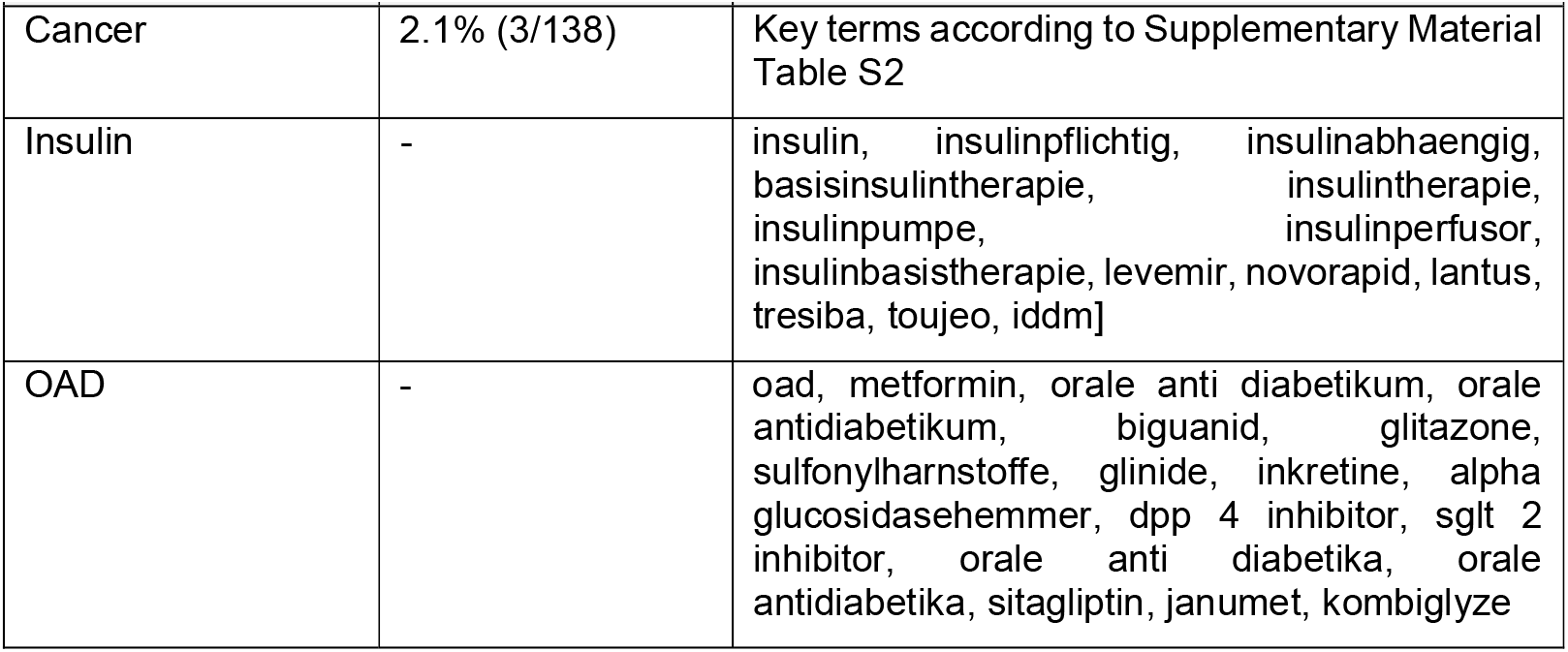
Overview on error rate of Natural Language Processing (NLP) detection of disease status on the validation dataset and the final key terms used for NLP.

**Table S2:**
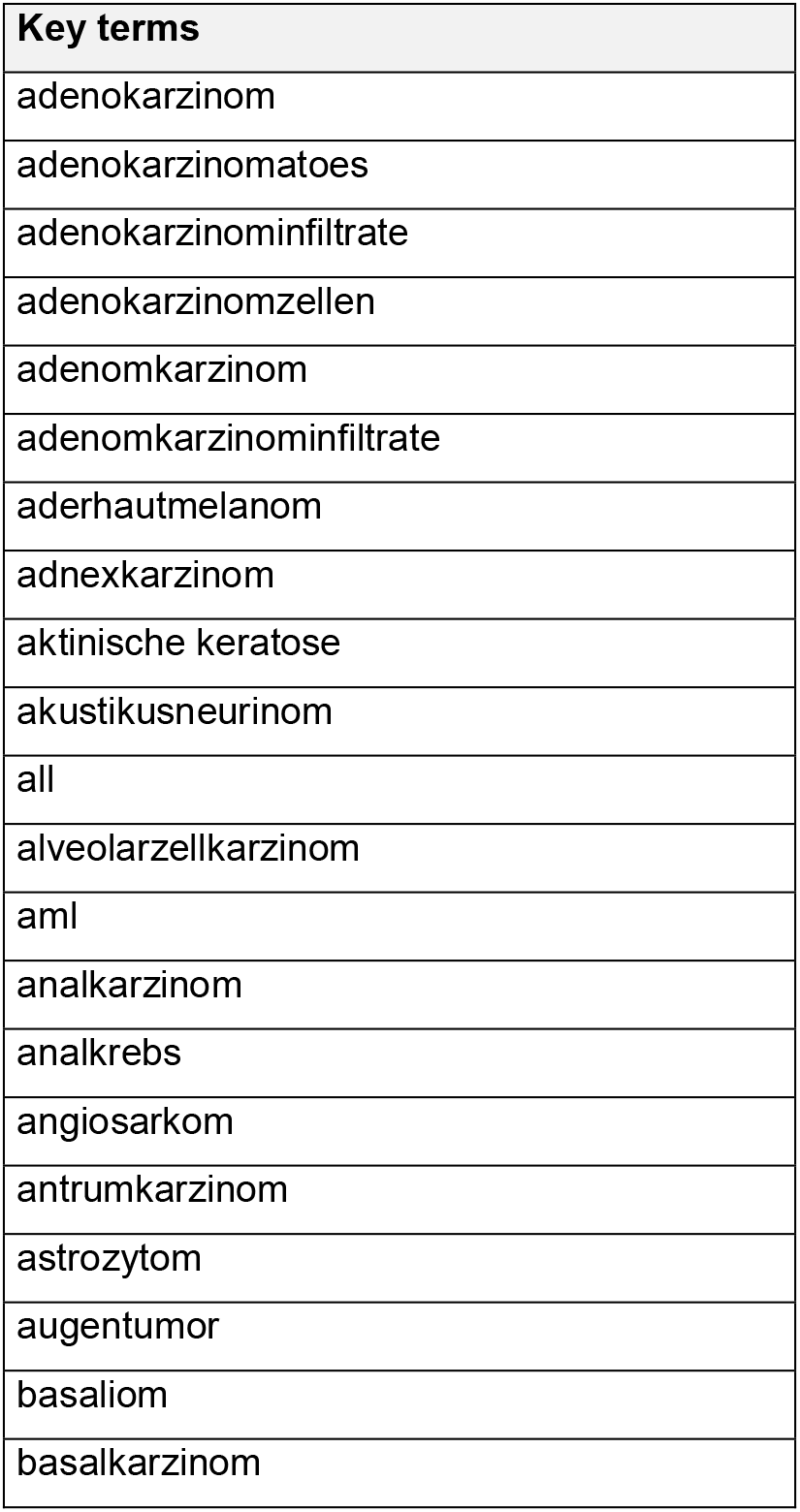
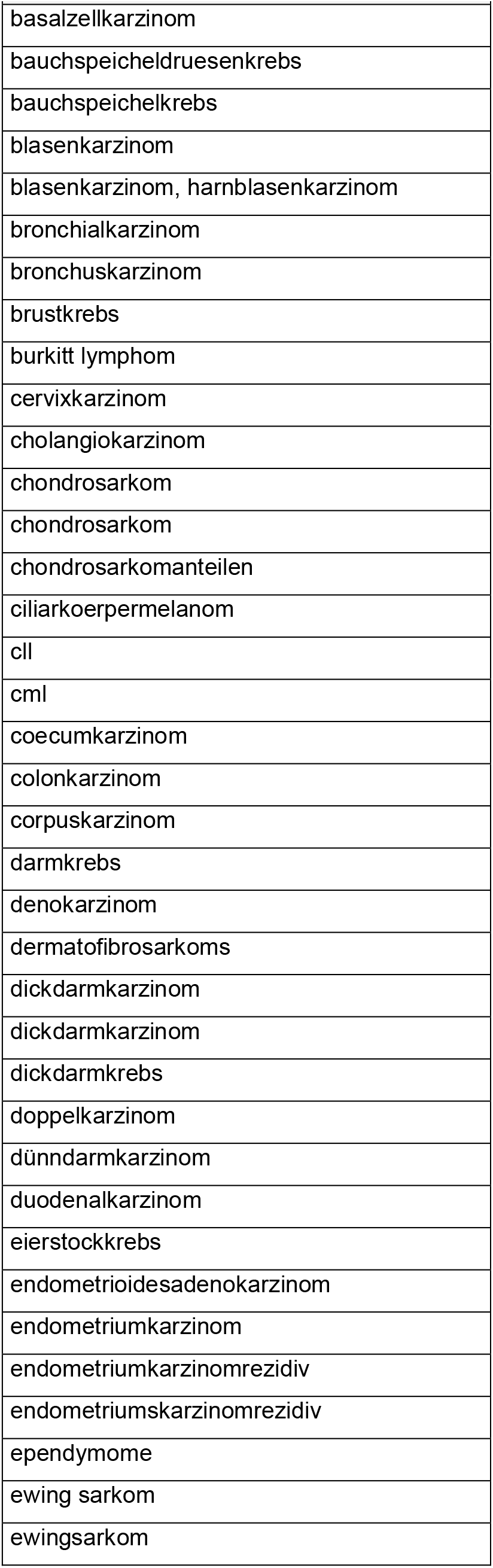
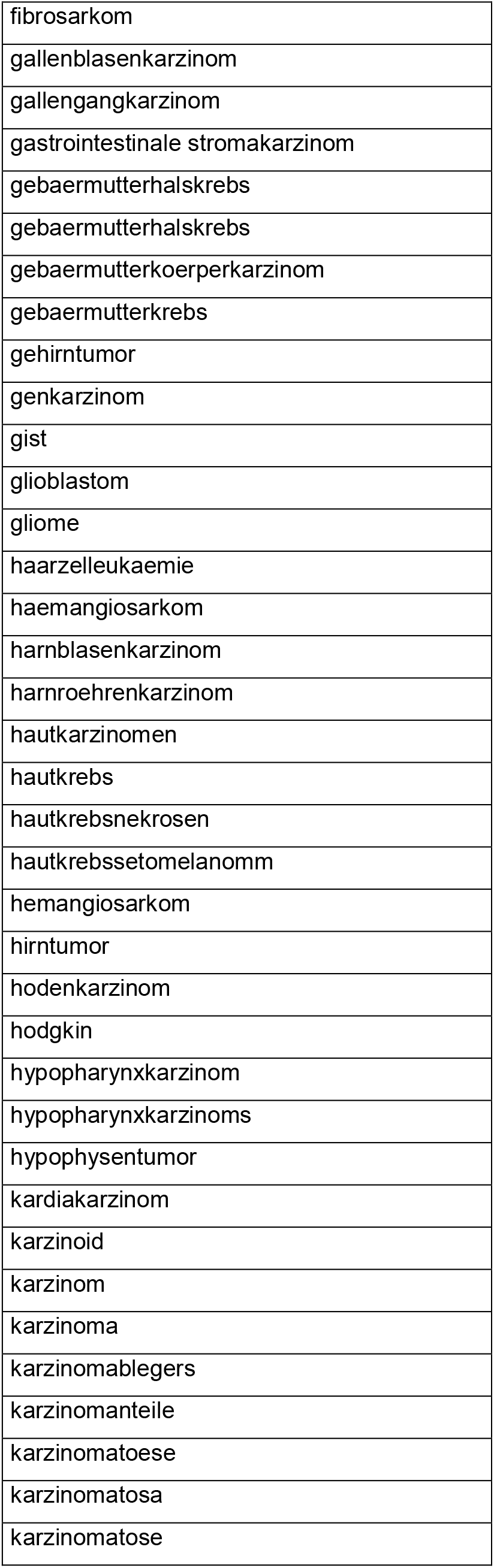
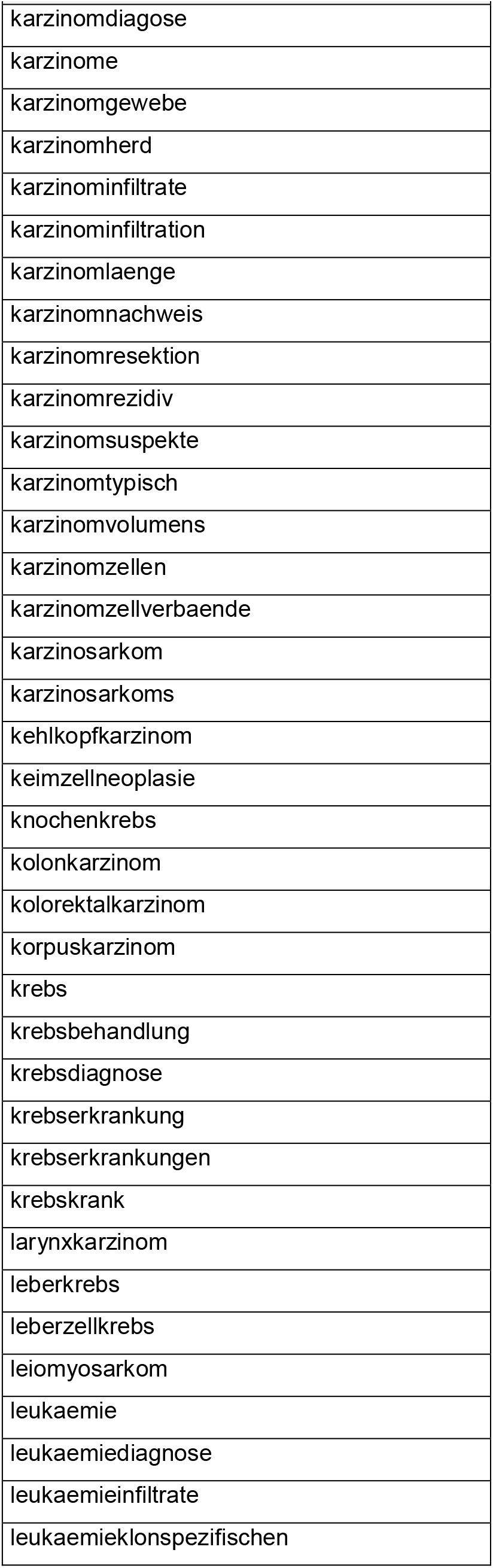
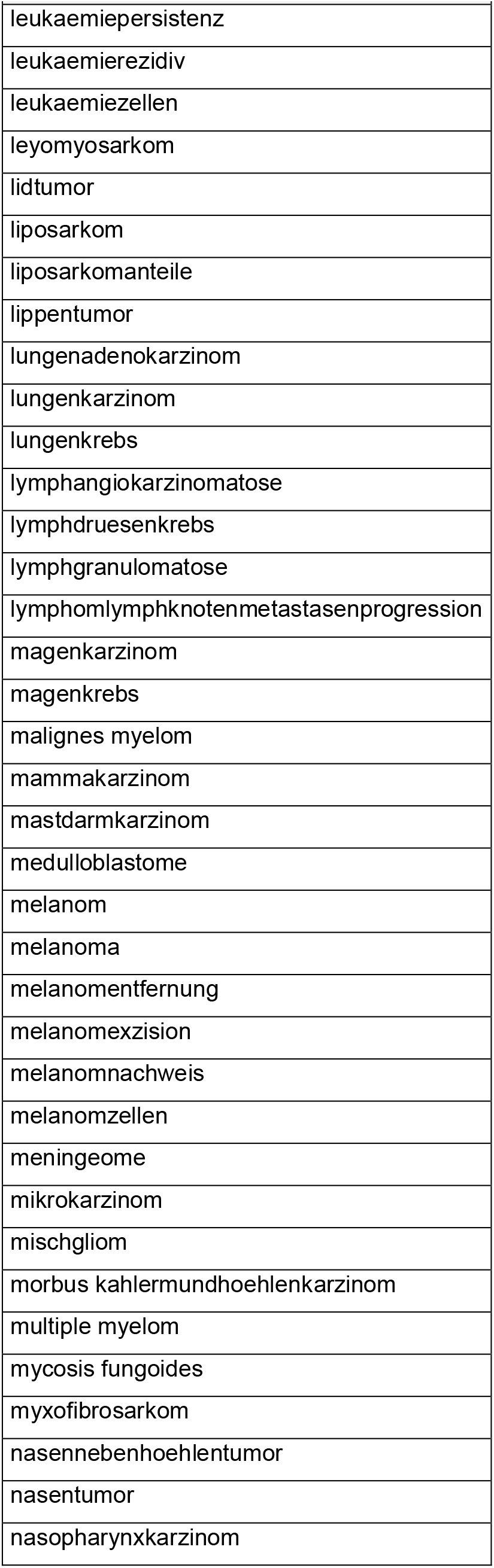
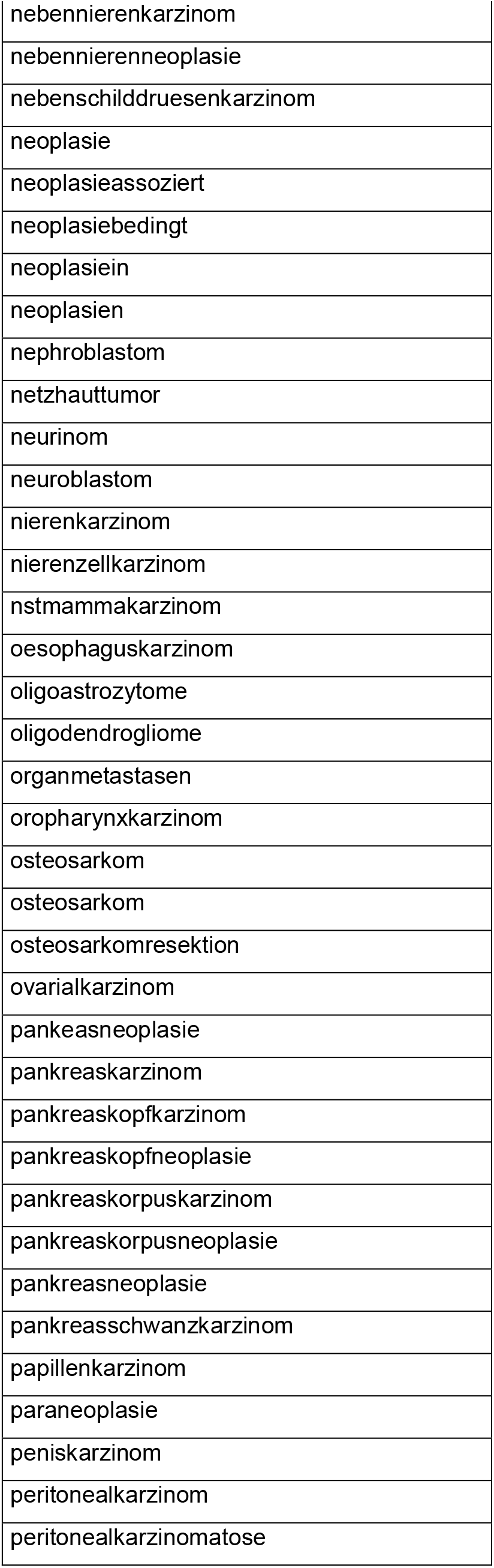
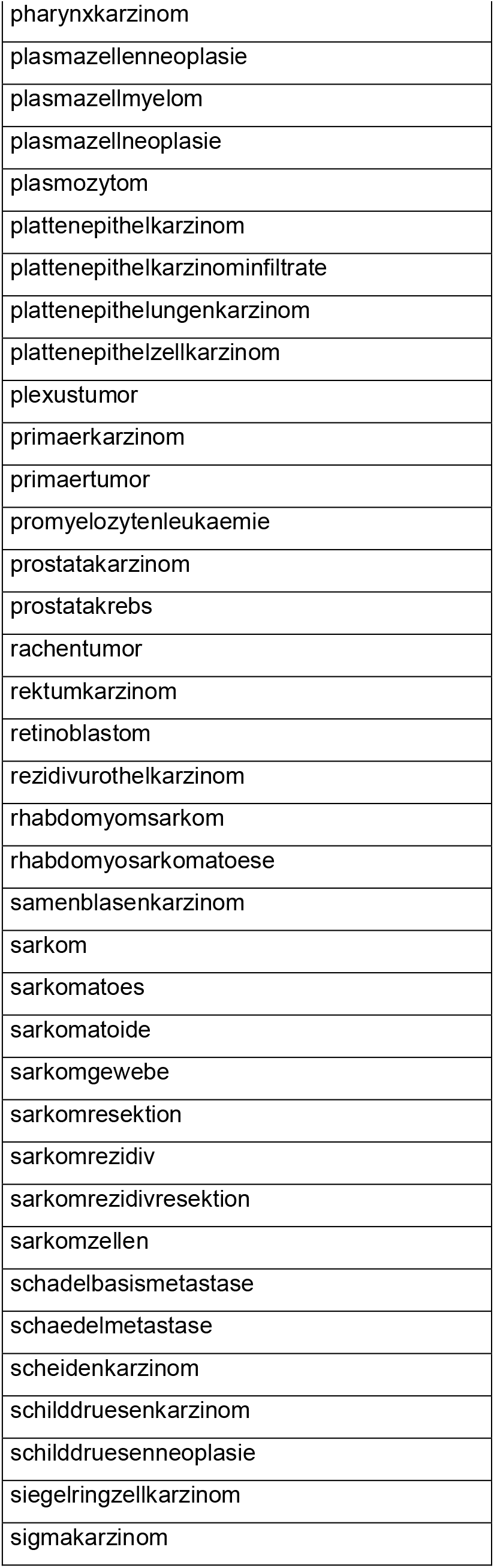
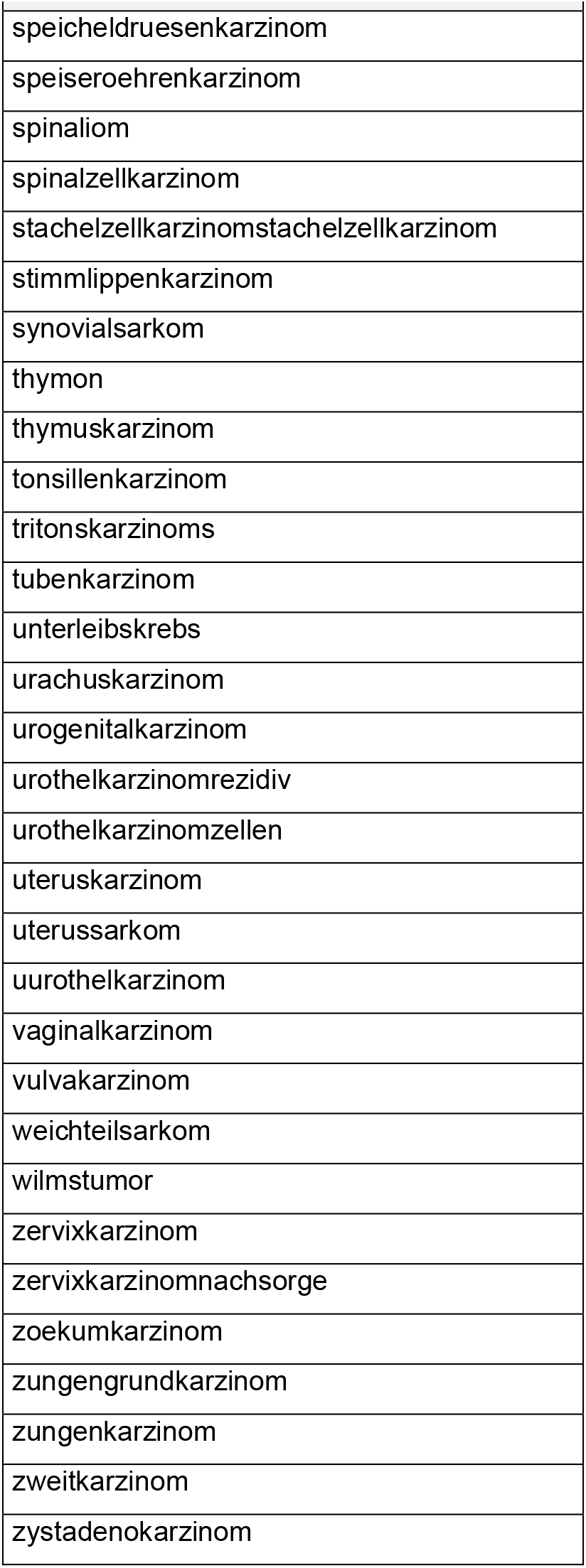
Key terms for the detection of cancer

## Notes

### Competing Interest Statement

The authors have declared no competing interest.

